# Adjudicating evolving inpatient diagnoses with a clinical coding agent: a retrospective multicentre study

**DOI:** 10.64898/2026.08.31.26361789

**Authors:** Yingda Li, Yinghong Shi, Yunfan Sun, Tao Suo, Peijia Wang, Zhou Zhang, Junli Xian, Lingqing Gu, Wei Ma, Jianying Gu, Jia Fan

**Affiliations:** Zhongshan Hospital, Fudan University, Shanghai, China; Mavcare AI Inc; Shidong Hospital of Yangpu District, Shanghai, China

## Abstract

**Background:** International Classification of Diseases (ICD) codes are widely used for reimbursement, health-service planning, and disease surveillance. Automated systems can identify diagnoses and supporting evidence across clinical documents. Yet coding is more than finding diagnoses. Decisions based on isolated information may not fully reflect the evolving clinical picture during admission. Provisional, superseded, or conflicting diagnoses can therefore persist in the final code set, complicating review and correction. We therefore developed and evaluated Clinico, a coding agent that organises evidence by time and clinical relationships to adjudicate diagnoses and update their status during admission.

**Methods:** We did a retrospective multicentre study with internal and external validation using inpatient records from two Shanghai hospitals. Routine coder-finalised codes served as references. Using 5,000 development stays, we refined Clinico’s prompts and workflow without fine-tuning model parameters. We compared Clinico with three prespecified comparators, two trained on 80,340 stays. Clinico maintained an evidence-linked diagnostic ledger across each admission and reconciled related or conflicting claims before finalising the code set and selecting the principal diagnosis. The primary outcome was principal-diagnosis exact-match agreement; complete code-set micro-F1 was the key secondary outcome.

**Findings:** Evaluation included 5,000 internal and 4,109 external test stays. Principal-diagnosis exact-match agreement was 83.6% (95% CI 82.6–84.6) internally and 52.0% (50.5–53.5) externally; the next-highest observed estimates were 74.7% and 31.5%. Complete code-set micro-F1 was 90.6% (95% CI 90.2–91.0) internally and 42.6% (42.0–43.2) externally; the next-highest observed estimates were 90.3% and 25.5%.

**Interpretation:** In both hospital test sets, Clinico had the highest observed estimates of principal-diagnosis agreement and complete code-set micro-F1 among evaluated methods. Its longitudinal adjudication process links decisions to source evidence, allowing them to be checked against the record. Prospective studies should assess its effects in clinical workflows using independently adjudicated references.

**Funding:** None.

## Introduction

The International Classification of Diseases (ICD) provides a common language for recording diagnoses in standardised categories, enabling coded health information to be compared across settings and over time.^1^ ICD-coded data support epidemiological surveillance, health-service planning, comparisons of hospital activity and quality, reimbursement, research, and comparisons of disease burden across populations.^2^ ICD is therefore a core component of modern clinical and administrative data infrastructure, but the value of this infrastructure depends on whether clinical episodes are represented consistently and faithfully.^3^

Producing such representations consistently and at scale is difficult. Even within a shared classification, outputs can vary with site-level coding and documentation practices and the interpretation of ambiguous records.^4,5^ Clinical information may be lost during clinical handovers,^6^ while interpretation of the record and its abstraction into codes introduce further opportunities for subjectivity, variability, and error.^4,7^ Within a hospital stay, information relevant to coding is often distributed across multiple clinical documents with variable structure and completeness.^8^ Working diagnoses may be revised, narrowed, or verified as evidence accumulates,^9^ and some test results may become available only after discharge.^10^ Many inpatient coding frameworks also require selection of a principal or main diagnosis, which under WHO ICD-10 guidance is the condition diagnosed at the end of the episode that was primarily responsible for the patient’s need for treatment or investigation.^11,12^ At hospital scale, documentation, clarification, code selection, and review span clinical and coding workflows.^4,13^ A systematic review estimated an average annual workload of about 11,300 records per professional coder in China. Under the study’s assumption of 220 eight-hour working days, this corresponds to about 9.3 min of available working time per record.^14^ Reliable automation cannot recover information that was never documented, but it could limit further information loss when the clinical episode is reviewed and coded. It could also support consistency within each framework, adaptation across settings, and efficient review without imposing identical coding across institutions.

Previous automated coding studies have addressed some of these tasks. Earlier systems such as PLM-ICD treated coding as document-level multilabel prediction from long clinical text.^8,15^ Later studies modelled document sequences across hospital stays,^16^ and recent workflows added diagnosis and evidence extraction, code retrieval, reranking, validation, and reconciliation.^17,18^ These methods can identify relevant information and select candidate codes. However, chronological ordering and candidate checking do not establish how the status of each diagnosis changes during care.^9,19,20^ Pathology reports may concern different specimens or sampling sites, and limited samples may support different levels of certainty. A provisional or less specific diagnosis may persist when definitive evidence is recorded separately or becomes available after discharge (appendix table A6). Chronology alone cannot determine whether labels describe the same condition, whether one should replace another, or whether both remain valid. Without this distinction, incompatible, duplicate, or outdated diagnoses may remain, or a supported diagnosis may be removed. A coding system must therefore link each diagnosis claim to source evidence and update its status. Related or conflicting claims must then be reconciled before the final code set and principal diagnosis are selected.

We therefore developed Clinico, an agent for inpatient ICD coding that reviews documents chronologically and maintains a source-linked diagnostic ledger. It updates claim status as evidence accumulates and reconciles related claims before finalising the code set and selecting the principal diagnosis. We compared Clinico with three prespecified comparators using internal and external test sets of routine inpatient records from two Shanghai hospitals. We separately explored its performance on Spanish CodiEsp cases.^21^

**Research in context**

**Evidence before this study**

We searched PubMed, ACL Anthology, arXiv, and medRxiv for studies published from Jan 1, 2011, to Aug 17, 2026, without language restrictions. Search terms combined ICD or clinical coding with machine learning, large language models, longitudinal or multi-document records, evidence extraction, diagnosis revision, adjudication, agentic workflows, and principal diagnosis. We included studies of automated ICD diagnosis coding from clinical records and excluded procedure-only coding, terminology mapping without records, and non-ICD classification. Earlier neural methods generally treated coding as document-level multilabel prediction. Later studies modelled temporal document sequences, updated code predictions during admission, linked codes to supporting text, and added retrieval, guideline application, verification, reconciliation, and principal-diagnosis selection. Most evidence came from retrospective single-centre studies or public benchmarks using routine codes as references. Few studies included external or prospective evaluation. Study heterogeneity precluded quantitative synthesis. Among the studies identified, we found no evaluation of an end-to-end inpatient coding system that maintained multiple source-linked diagnosis claims across chronologically reviewed documents, revised their status as evidence accumulated, and used this longitudinal state to reconcile the final diagnosis set and select the principal diagnosis.

**Added value of this study**

This study evaluated Clinico, a coding agent that treated inpatient diagnosis coding as longitudinal adjudication. It reviewed documents chronologically and maintained multiple diagnosis claims, their evolving status, clinical relationships, and source evidence. Claims were revised and reconciled before the diagnosis-code set was finalised and the principal diagnosis selected. Without task-specific parameter fine-tuning, Clinico had the highest observed complete code-set micro-F1 and principal-diagnosis exact-match agreement among three prespecified comparators in both hospital test sets. Component analyses showed lower principal-diagnosis agreement when pathology-report processing or diagnosis reconciliation was omitted, or when the cumulative hospital-course account was replaced by per-claim evidence summaries. Final decisions remained linked to source evidence. A separate exploratory analysis assessed Spanish clinical text and a different coding system.

**Implications of all the available evidence**

These findings support longitudinal diagnosis-state adjudication as a promising direction for automated inpatient coding. Prospective evaluation is needed to establish clinical effectiveness and support broader deployment. Agreement with routine coder-finalised codes does not establish accuracy against an independently adjudicated reference. Source-linked outputs were designed for human review, but effects on error correction, review time, usability, and professional judgement were not established. Prospective multicentre human-in-the-loop studies should evaluate these outcomes across hospitals and coding systems using independently adjudicated references. If confirmed, this approach could support coding review and quality assurance and improve the reliability of data used for reimbursement, surveillance, benchmarking, registries, research, and health-service planning.

## Methods

### Study design and setting

We did a retrospective multicentre development and validation study of Clinico using routinely collected inpatient electronic health records from Zhongshan Hospital, Fudan University, and Shidong Hospital of Yangpu District, Shanghai, China. Zhongshan supported development and internal testing, Shidong external testing, and CodiEsp exploratory Spanish-language benchmarking. Ethics approvals (Zhongshan B2026-078; Shidong 2026-017-01) included consent waivers; direct identifiers were removed before export.

### Data sources and participants

The analysis unit was a stay from admission to discharge. We screened all stays without diagnosis-based or department-based sampling, excluding those missing an admission note, discharge summary, or coder-reviewed final code set. We retained each patient’s most recent eligible stay and ordered documents chronologically.

Zhongshan, a tertiary referral hospital, included 90,340 stays from May 2025 to January 2026, randomly divided into comparator training (80,340), development (5,000), and internal testing (5,000). Shidong, a secondary district hospital, contributed 4,109 stays from May to July 2025 for external testing.

Documents included admission, progress, discharge, procedure, and available pathology records. Structured fields provided age (final digit masked before analysis), recorded sex, admission department, and admission and discharge dates. Zhongshan discharge summaries commonly contained unstandardised free-text diagnosis sections. Unlike reference labels, these inputs could omit diagnoses, vary in terminology or specificity, or predate pathology. They were not independently adjudicated. At Shidong, diagnosis sections and some document types were inconsistently available. Dates within each stay were shifted by a common random offset, preserving order and intervals.

We separately analysed CodiEsp’s 250-case held-out set in Spanish.^21^

### Input preparation

Before development and evaluation, entries in each clinician-authored diagnosis section were randomly reordered once with the same permutation across methods and runs. Content, chronology, and reference codes were unchanged.

### Reference labels

Hospital reference labels were structured codes finalised through routine professional review of documentation and linked pathology, including post-discharge reports, under Chinese ICD-10-based procedures. Zhongshan used its ordered administrative code list and Shidong the union of its administrative and clinical lists. Newly linked postdischarge pathology prompted updates to diagnosis, code, and principal-diagnosis suggestions for clinician and coder review. Under national rules, the first code was principal and the remainder secondary; their order was not analysed.^13^ All codes formed the complete-set reference. Labels were not independently recoded or clinically adjudicated. CodiEsp labels were the dataset’s CIE-10-ES diagnosis-code annotations.^21^

All reference labels predated Clinico deployment and were not informed by its outputs.

### Clinico

Clinico was a stateful coding agent maintaining a persistent diagnostic ledger per stay (figure 1). Non-pathology documents were processed chronologically. For each, diagnosis review used the ledger to extract new or revised claims and assign *add, confirm, refine, correct*, or *negate* actions; separate evidence review linked outputs to dated source snippets. Claims formed longitudinal chains; nodes sharing a standardised diagnosis and code were merged, retaining provenance and relations.

**Figure 1:**
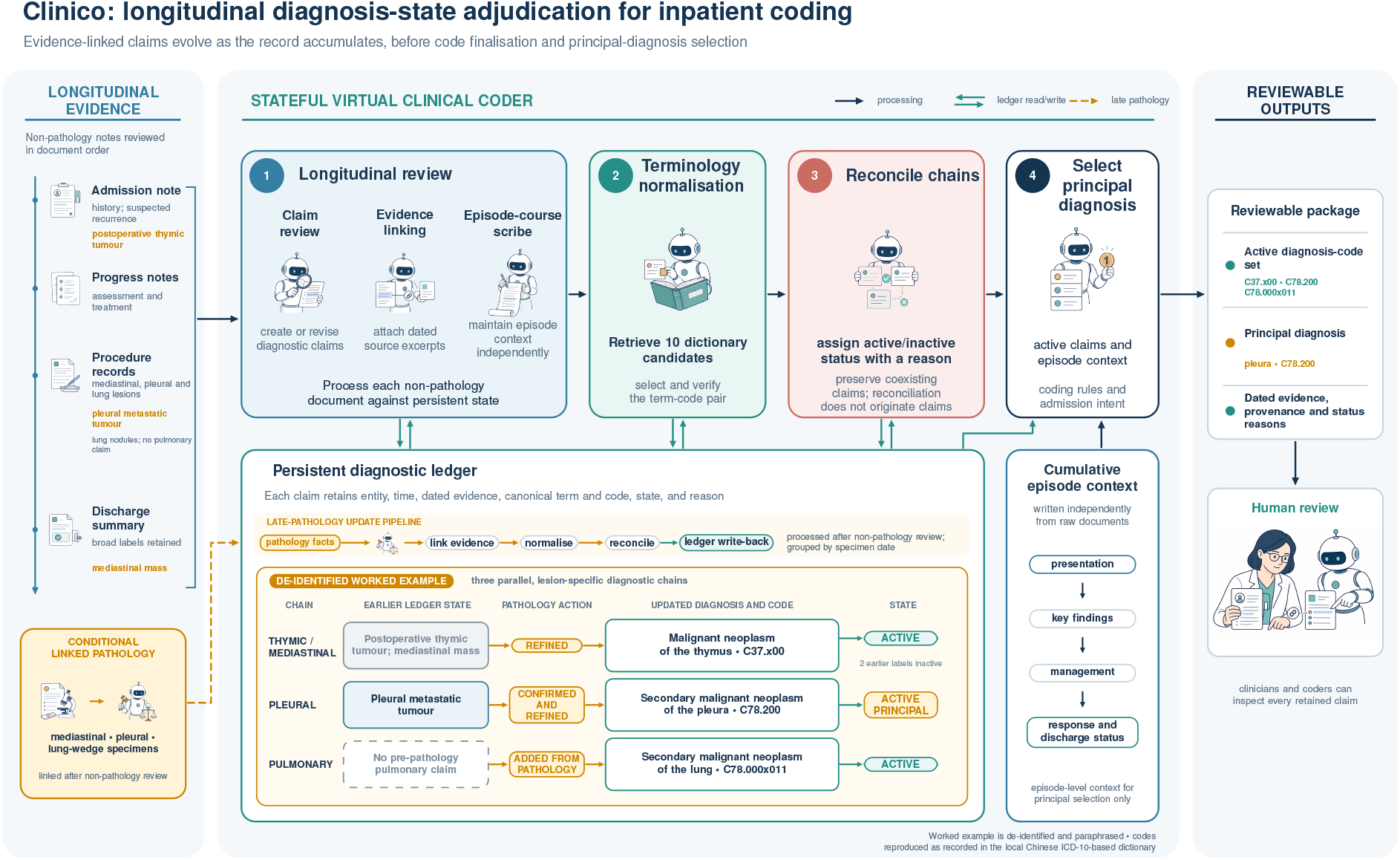
Clinico architecture for longitudinal diagnosis-state adjudication. Non-pathology documents are reviewed in document order while diagnosis claims, dated evidence, canonical terms, codes, status, and reasons are retained in a persistent ledger. Linked pathology is processed after non-pathology review and can refine, confirm, or add lesion-specific claims. Reconciled active claims and cumulative episode context support principal-diagnosis selection; retained provenance permits clinician and coder review. The embedded de-identified worked example shows three coexisting lesion-specific diagnostic chains.

Each claim was canonicalised against the diagnosis dictionary: the embedding model retrieved ten candidates and the language model selected a diagnosis and code from stored evidence. A verifier selected another retrieved candidate or rejected mappings flagged by predefined similarity or ambiguity rules. Rejected claims were retained inactive with a recorded reason and excluded; accepted claims remained active until reconciliation.

Linked pathology reports were then grouped by specimen date and processed sequentially with the complete ledger. For the same lesion, an explicit conclusion outweighed earlier statements but did not override unrelated claims. Without an explicit conclusion, morphology and immunohistochemistry could generate or revise candidates, which underwent evidence linking, canonicalisation, and reconciliation without independent expert review.

Claim evidence summaries used only stored source snippets. Chains were reconciled separately using action histories and summaries. Reconciliation assigned active or inactive status with a reason but could not create diagnoses, change standardised diagnoses or codes, or link chains. Coexisting diagnoses could remain active, and all active, canonicalised claims formed the final code set.

Finally, Clinico used a cumulative clinical-course summary to select the principal diagnosis from active diagnoses and provide a reason.

A de-identified worked example is provided in the appendix (figure A1).

Language-model calls used Qwen3-32B-FP8 and retrieval Qwen3-Embedding-4B.^22,23^ Both ran on private servers. Neither was fine-tuned on study records. Prompts and workflow settings were refined on Zhongshan development data and frozen before testing.

### Comparator systems

Comparators were fixed before testing. PLM-ICD followed its published document-level formulation and used discharge summaries. LLM-ICD also received discharge summaries because a full record could exceed the context of one model call. The adapted MedCodER workflow received all available documents. All methods received complete CodiEsp reports.

PLM-ICD retained the published segment-pooling and label-aware-attention architecture.^15^ Separate models independently predicted the complete set and principal diagnosis; the latter was not added to the complete set, and outputs were restricted to training codes. Models were fitted separately to Zhongshan and official CodiEsp training data with corresponding development-set selection. The hospital complete-set threshold maximised micro-F1 over a prespecified 0.01–0.99 Zhongshan development-set grid and was fixed before testing.

LLM-ICD used Qwen3-32B-FP8 with separate low-rank adaptations fitted on Zhongshan and CodiEsp training sets.^22,24^ It generated ordered sets; the first hospital code was principal.

The adapted workflow processed documents chronologically, extracted diagnoses and evidence, and retrieved and reranked codes; overlength inputs were truncated.^17^ A study-defined two-stage hospital selector classified admission intent and selected the principal diagnosis from the final set. It did not receive Clinico’s cumulative summary and could not create diagnoses.

Qwen methods shared checkpoint, precision, context length, and decoding settings. Hospital configurations and the shared Chinese diagnosis dictionary were fixed using Zhongshan training and development data before Shidong testing. No test data informed fitting or tuning; Clinico and MedCodER were not fitted to CodiEsp.

### Outcomes and statistical analysis

The primary outcome was principal-diagnosis exact-match agreement with the routine hospital reference; missing, invalid, or unmapped outputs were incorrect. CodiEsp lacks principal labels. Complete-set secondary outcomes were micro-precision, micro-recall, micro-F1, and macro-F1. Micro metrics pooled true-positive, false-positive, and falsenegative codes across records. Hospital macro-F1 used the union of reference and aligned predicted labels for each method or configuration, with unmatched predictions retained as zero-F1 labels. CodiEsp used a fixed reference-code set across methods.

Predictions were compared by exact identifiers; unmapped predictions remained errors. Shidong reference codes were seen if the exact identifier occurred in Zhongshan training and unseen otherwise. We calculated 95% CIs from 10,000 non-parametric bootstrap resamples by hospital stay or CodiEsp case, unless otherwise stated. Between-method differences were reported in percentage points without CIs. All eligible records were included without samplesize calculation. Reporting followed STROBE and RECORD.^25,26^

### Post-hoc component ablation analysis

In the Zhongshan internal test set, post-hoc analyses separately omitted pathology-report processing and reconciliation, replaced the cumulative summary with per-claim evidence summaries, and combined these modifications. Other inputs, models, dictionaries, and inference settings were fixed. We reported absolute performance and paired full-minus-ablated differences with 95% CIs from 10,000 stay-level bootstrap resamples. Configuration details are in the appendix.

### Exploratory computational throughput analysis

Using four RTX 4090D GPUs, we ran the frozen workflow end to end on 100 randomly sampled Zhongshan test stays. Timing excluded model and service start-up and a separate warm-up stay. We reported throughput and amortised time per stay, not single-stay latency; no CI was calculated. Full settings are in the appendix.

### Exploratory physician survey

We did an exploratory post-implementation survey at Zhongshan Hospital on July 20–24, 2026, during operational use of Clinico for inpatient discharge-abstract completion. A voluntary Chinese-language online questionnaire was distributed to physicians in 40 inpatient departments. The number receiving or viewing the invitation was unavailable, precluding an individual response proportion. Responses were summarised as counts and percentages. The committee classified this as an internal service evaluation not requiring research ethics review.

## Results

At Zhongshan, 105,713 hospital stays were screened and 15,373 excluded after eligibility assessment and retention of the most recent eligible stay per patient, leaving 90,340: 80,340 for PLM-ICD and LLM-ICD training and 5,000 each for internal development and testing. Shidong contributed 4,109 stays in the external test set, giving 94,449 hospital stays in the final analytic datasets. Shidong stays contained less than half as much text as Zhongshan test stays but more distinct reference diagnosis codes (median 5 vs 4 per stay). Circulatory-system codes were most frequent in both test sets, while Zhongshan had more Z and malignant-neoplasm codes and Shidong more digestive-system and endocrine or metabolic codes.

At the full-code level, 7,621 (36.0%) of 21,159 assignments and 665 (38.1%) of 1,747 unique Shidong codes had not been observed in Zhongshan training data (table 1).

**Table 1:**
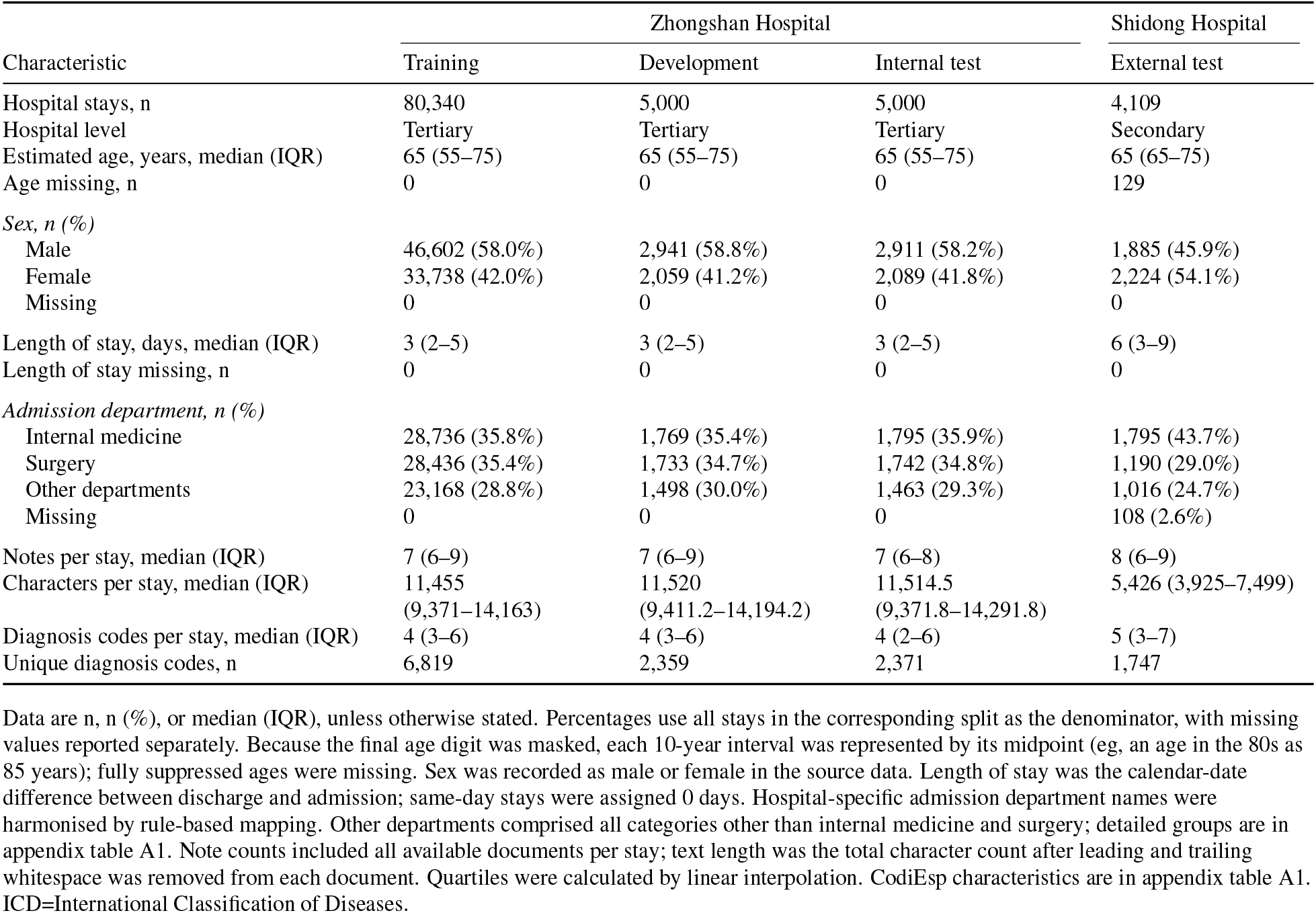
Characteristics of hospital analytic datasets.

Clinico had the highest observed point estimate for principal-diagnosis exact-match agreement in both hospital test sets (table 2; figure 2). Agreement was 83.6% (95% CI 82.6–84.6) in the Zhongshan internal test set; the point estimate was 8.9 percentage points higher than that of PLM-ICD (74.7% [73.5–75.9]), which had the next-highest observed estimate. In the Shidong external test set, agreement was 52.0% (50.5–53.5), compared with the next-highest observed estimate of 31.5% (30.1–32.9) for MedCodER.

**Table 2:** Performance of Clinico and prespecified comparators in hospital test sets.

| Test set | Method | Principal diagnosis agreement | Micro-F1 | Micro-precision | Micro-recall | Macro-F1 |
| --- | --- | --- | --- | --- | --- | --- |
| Zhongshan<br>Internal test | PLM-ICD | 74.7%<br>(73.5–75.9) | 36.6%<br>(35.9–37.4) | 60.4%<br>(59.5–61.3) | 26.3%<br>(25.6–27.0) | 1.0%<br>(1.0–1.1) |
|  | LLM-ICD | 71.2%<br>(70.0–72.5) | 90.3%<br>(89.8–90.7) | 91.5%<br>(91.0–92.0) | 89.1%<br>(88.5–89.7) | 68.6%<br>(67.2–69.9) |
|  | MedCodER | 54.8%<br>(53.4–56.1) | 78.6%<br>(78.1–79.1) | 74.3%<br>(73.7–74.9) | 83.3%<br>(82.7–83.9) | 54.6%<br>(53.5–55.7) |
|  | Clinico | 83.6%<br>(82.6–84.6) | 90.6%<br>(90.2–91.0) | 89.5%<br>(89.0–90.0) | 91.8%<br>(91.3–92.2) | 72.6%<br>(71.2–74.1) |
| Shidong<br>External test | PLM-ICD | 13.5%<br>(12.5–14.6) | 0.2%<br>(0.1–0.2) | 25.8%<br>(15.3–37.3) | 0.1%<br>(0.0–0.1) | 0.03%<br>(0.00–0.05) |
|  | LLM-ICD | 15.7%<br>(CI pending) | 16.9%<br>(CI pending) | 28.1%<br>(CI pending) | 12.1%<br>(CI pending) | 9.6%<br>(9.0–10.2) |
|  | MedCodER | 31.5%<br>(30.1–32.9) | 25.5%<br>(24.9–26.0) | 27.0%<br>(26.4–27.7) | 24.1%<br>(23.5–24.7) | 14.5%<br>(14.0–15.0) |
|  | Clinico | 52.0%<br>(50.5–53.5) | 42.6%<br>(42.0–43.2) | 37.8%<br>(37.2–38.5) | 48.8%<br>(48.1–49.5) | 24.4%<br>(23.7–25.0) |

**Figure 2:**
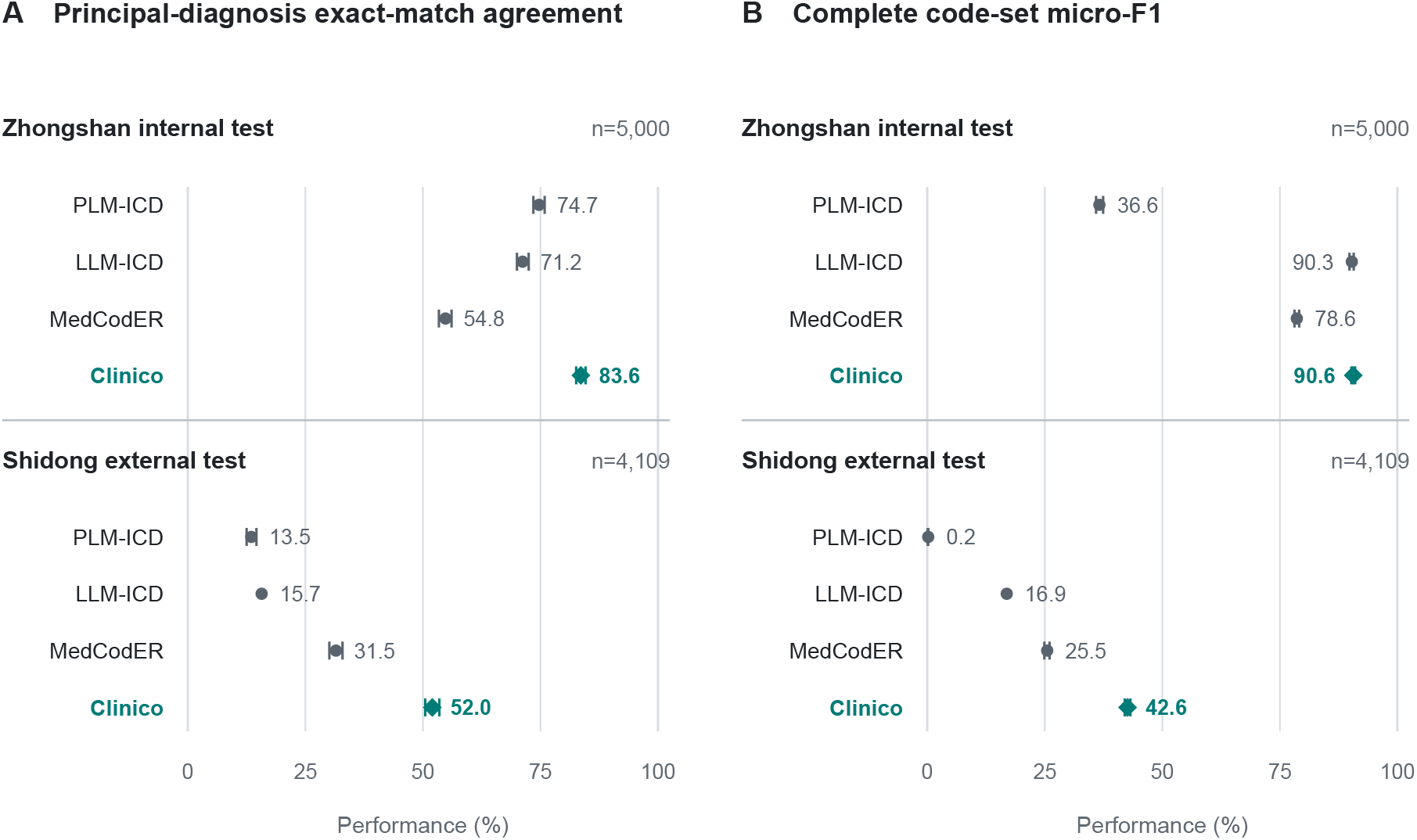
Performance in the Zhongshan internal and Shidong external hospital test sets. (A) Principal-diagnosis exact-match agreement. (B) Complete diagnosis-code-set micro-F1. Points show estimates and horizontal lines show available 95% CIs. CIs for the corrected full-cohort Shidong LLM-ICD estimates await recalculation and are not shown. Shidong evaluation was done without Shidong-data-informed model fitting or hyperparameter tuning. For PLM-ICD, principal-diagnosis and complete code-set outcomes were produced by separately trained models and evaluated as separate outputs. CIs describe within-method estimates, not between-method differences. ICD=International Classification of Diseases. LLM=large language model.

For the complete diagnosis set at Zhongshan, Clinico had a micro-F1 of 90.6% (95% CI 90.2–91.0), microprecision of 89.5% (89.0–90.0), micro-recall of 91.8% (91.3–92.2), and macro-F1 of 72.6% (71.2–74.1). Its micro-F1 was 0.4 percentage points above LLM-ICD (90.3% [89.8–90.7]), which had the highest observed micro-precision; Clinico had the highest observed micro-recall and macro-F1.

All methods were applied to the Shidong external test set without Shidong-data-informed model fitting or hyperparameter tuning. Clinico’s external micro-F1 was 42.6% (95% CI 42.0–43.2), compared with the next-highest observed estimate of 25.5% (24.9–26.0) for MedCodER. Clinico’s micro-precision was 37.8% (37.2–38.5), micro-recall was 48.8% (48.1–49.5), and macro-F1 under the legacy diagnosis-name metric was 24.4% (23.7–25.0). MedCodER had higher observed point estimates than both Zhongshan-trained comparators for external principal-diagnosis agreement and micro-F1. At the development-selected threshold of 0.28, the PLM-ICD complete-set model generated 62 predictions across 43 of 4,109 stays, of which 16 matched reference codes; micro-F1 was 0.2% (0.1–0.2), micro-precision was 25.8% (15.3–37.3), and micro-recall was 0.1% (0.0–0.1). Detailed results are shown in table 2.

In the post-hoc component ablation analysis in the Zhongshan internal test set, omission of pathology-report processing or diagnosis reconciliation, or replacement of the cumulative clinical-course summary with per-claim evidence summaries, reduced principal-diagnosis agreement by 7.5 percentage points (95% CI 6.6–8.5), 5.6 points (4.8–6.5), and 11.0 points (9.9–12.0), respectively, compared with the full workflow; joint omission of all three components reduced agreement by 15.0 points (13.8–16.3). Omitting pathology-report processing or diagnosis reconciliation increased micro-recall by 1.2 and 2.8 points, respectively, but reduced micro-precision by 2.5 and 7.0 points, micro-F1 by 0.7 and 2.5 points, and macro-F1 by 1.0 and 5.0 points. When the cumulative clinical-course summary was replaced, the complete diagnosis set was unchanged by design and the diagnosis-set metrics were therefore identical to those of the full workflow. Joint omission increased micro-recall by 1.6 points but reduced micro-precision, micro-F1, and macro-F1 by 5.3, 2.1, and 2.6 points, respectively (appendix tables A4 and A5).

The CodiEsp test set included 250 curated Spanish clinical cases. Because CodiEsp did not provide a principal-diagnosis reference label, evaluation was limited to complete diagnosis-code set performance. Clinico had a micro-F1 of 31.7% (95% CI 29.5–33.8), micro-precision of 79.2% (76.2–82.1), micro-recall of 19.8% (18.2–21.4), and macro-F1 of 27.2% (25.5–29.0). Its micro-F1 point estimate was 4.3 percentage points lower than that of PLM-ICD (36.0% [34.0–38.1]) and was the second-highest observed point estimate. Clinico had the highest observed micro-precision, whereas MedCodER had the highest observed macro-F1 (28.5% [26.6–30.4]). Full CodiEsp results are shown in table 3.

**Table 3:** Performance on CodiEsp Spanish source texts.

| Method | Micro-F1 | Micro-precision | Micro-recall | Macro-F1 |
| --- | --- | --- | --- | --- |
| PLM-ICD | 36.0%<br>(34.0–38.1) | 54.3%<br>(50.7–58.2) | 27.0%<br>(25.0–29.0) | 7.2%<br>(6.6–7.8) |
| LLM-ICD | 23.4%<br>(21.1–25.7) | 25.5%<br>(23.1–27.8) | 21.7%<br>(18.7–24.7) | 11.8%<br>(10.7–12.9) |
| MedCodER | 30.6%<br>(28.3–32.9) | 65.5%<br>(62.1–68.8) | 20.0%<br>(18.2–21.8) | 28.5%<br>(26.6–30.4) |
| Clinico | 31.7%<br>(29.5–33.8) | 79.2%<br>(76.2–82.1) | 19.8%<br>(18.2–21.4) | 27.2%<br>(25.5–29.0) |

Data are percentages with 95% CIs, unless otherwise stated. CodiEsp was evaluated using Spanish source texts. Predictions were compared using exact full-code identifiers after lowercasing and trimming surrounding whitespace; invalid or unmapped predictions were retained as errors. Micro-F1, micro-precision, and micro-recall were calculated for the complete diagnosis-code set. Macro-F1 used the fixed set of reference codes observed in the complete test set, shared across methods. CIs were derived from 10,000 case-level bootstrap resamples; percentile intervals were used for micro metrics, and the macro-F1 interval was calculated from the bootstrap standard error and centred on the full-sample estimate. Between-method differences are descriptive, were calculated from unrounded point estimates, and do not have paired difference CIs. Principal-diagnosis agreement was not assessed because CodiEsp did not provide a principal-diagnosis reference label. LLM=large language model.

Clinico processed all 100 sampled stays in 232.2 min (25.8 stays per h; amortised wall-clock time 2.322 min per stay), without GPU out-of-memory events (appendix table A2).

In the separate exploratory post-implementation survey, 64 physicians from 24 of 40 inpatient departments completed the questionnaire (departmental coverage 60.0%). Of these, 33 (51.6%) considered the coding analyses generated by artificial intelligence (AI) reasonable, and 22 (34.4%) reported never adopting an AI analysis to revise a code. Thirty-eight (59.4%) perceived an improvement in coding quality within the medical-records department after implementation. Forty-nine (76.6%) respondents considered the analyses helpful in promoting standardised documentation in subsequent inpatient discharge abstracts, and 51 (79.7%) considered them useful for providing educational guidance on subsequent coding (appendix table A3). These item-level percentages use all 64 voluntary respondents as the denominator and are not estimates for all physicians at the hospital.

## Discussion

Clinico had the highest observed principal-diagnosis agreement and complete code-set micro-F1 in both hospital test sets. Clinico’s internal micro-F1 was 90.6% without task-specific parameter fitting, compared with 90.3% for LLM-ICD after low-rank fine-tuning. Clinico retained the highest observed estimates after external transfer to Shidong. These findings support longitudinal diagnostic discovery and adjudication as a distinct formulation of inpatient coding.

Earlier multilabel models predicted codes from clinical text, and later work modelled temporal document sequences across a hospital stay; other workflows added extraction, retrieval, reranking, verification, and guideline reconciliation.^15,16,17,18^ Clinico addresses an upstream question by tracking diagnoses as source-linked claims that may be confirmed, refined, replaced, or excluded as evidence accumulates. Discovery and final retention were therefore separate steps; removing reconciliation increased recall but reduced precision, micro-F1 by 2.5 points, and macro-F1 by 5.0 points. Reconciliation improved agreement by producing a coherent final set, rather than simply retaining more candidates. This distinction matters clinically because information is often repeated across clinical documents, while diagnoses can be narrowed or replaced as evidence accumulates.^27,9^

Pathology could become available after the discharge summary. Clinico could infer a diagnostic candidate from morphology and immunohistochemistry when the report lacked a definitive conclusion. After canonicalisation and reconciliation, that diagnosis could enter the final set or become principal; newly linked post-discharge pathology also triggered automatic updates in operational use. Professional coders reviewed and corrected these outputs before final entry, reflecting the local workflow in which later pathology could revise an initial diagnosis.^10,9^ The ablation pattern was consistent with pathology supporting selective refinement rather than indiscriminate expansion of the diagnosis set. Retrospective pathology-derived inferences were not separately adjudicated by pathologists, making targeted expert review an important next evaluation.

Principal-diagnosis selection provided complementary evidence of episode understanding: replacing the cumulative hospital-course summary with per-claim evidence summaries left the diagnosis set unchanged but reduced agreement by 11.0 percentage points. The integrated account therefore provided information beyond the evidence summaries for individual diagnoses.

PLM-ICD and LLM-ICD were trained on 80,340 Zhongshan stays, whereas Clinico did not fit parameters to those labelled stays. Clinico used local development data to refine prompts and workflow and configure the diagnosis dictionary. Its comparable internal performance shows that strong results were possible without task-specific parameter fitting. MedCodER shared the language and embedding models and received all available documents, making it the most informative workflow comparator. Clinico’s higher observed estimates in both hospitals are consistent with added value from longitudinal claims, reconciliation, and hospital-course reconstruction. The workflows also differed in truncation, canonicalisation, and principal-diagnosis selection, so the comparison supports the integrated formulation rather than one isolated component.

Shidong was a clinically meaningful stress test with less complete and less standardised data. Records contained half the text of Zhongshan records, and document availability was inconsistent. In addition, 36.0% of reference assignments and 38.1% of unique codes were unseen in Zhongshan training data. Some reference codes were more specific than available documentation (eg, rhinovirus-associated acute bronchitis when records supported acute bronchitis but did not identify rhinovirus). Combination diagnoses were also more common; ureteric calculus, hydronephrosis, and infection could be recorded separately but represented by one code. Some labels lacked identifiable support in exported documents, possibly reflecting indirect documentation, information available to coders but absent from model inputs, or routine coding differences. Clinico retained the highest observed principal-diagnosis agreement and micro-F1, although data limitations and model errors constrained absolute agreement.

The operational workflow was human-in-the-loop: clinicians and coders could inspect suggested codes, evidence, the hospital course, and diagnostic evolution. Related studies have evaluated interactive coder support and real-world human–AI coding workflows.^28,29^ Professional coders retained final responsibility, while about four in five survey respondents reported educational or standardisation value. However, 51.6% considered the analyses reasonable and 34.4% never adopted a suggestion. Users may therefore value evidence organisation without accepting every recommendation, a hypothesis that prospective studies should assess against objective coding outcomes.

CodiEsp provided a separate test of modular adaptation. Clinico achieved the highest observed precision without task-specific parameter fitting, although recall was lower. Its 250 single texts and absence of principal labels^21^ make it a test of language and terminology adaptation rather than longitudinal reasoning. Batch processing was technically feasible on the tested four-GPU configuration, while clinical efficiency still requires end-to-end human-in-the-loop evaluation.

Several considerations define the scope. Hospital references predated Clinico deployment and were not informed by its outputs, but they were not independently clinically adjudicated; routine discharge-code accuracy varies against independent record review.^3^ Exact-code agreement does not validate intermediate reasoning; between-method differences were descriptive, and comparators differed in training, inputs, and output constraints. External validation involved one additional hospital in the same city, without subgroup analyses of documentation completeness, specialty, or case complexity. The next priorities are blinded adjudication, broader external validation, and prospective evaluation of coding quality and workflow.

Clinico extends one-time code prediction to updateable diagnostic-state adjudication. The immediate opportunity is dynamic revision when late evidence arrives, with quality control directing attention to unsupported or consequential diagnostic changes. An inspectable layer linking raw records, evolving claims, professional review, and standardised codes could extend this framework beyond coding, provided each use is validated. Preserving diagnostic evolution can organise fragmented evidence for adjudication and final coding.

## Data Availability

The individual-level hospital records and survey responses are not publicly available because they contain potentially sensitive information and are subject to institutional data-governance and ethics restrictions. Requests for access may be submitted to the corresponding authors and will be considered by the participating institutions and relevant ethics committees; access cannot be guaranteed. Aggregate data supporting the findings are provided in the manuscript and appendix. The CodiEsp dataset is publicly available at https://doi.org/10.5281/zenodo.3837305.

## Appendix

### Detailed dataset characteristics

appendix table A1 reports the complete demographic, admission-department, document, and coding characteristics of the analytic datasets.

**Table A1:** Detailed characteristics of analytic datasets.

| Characteristic | Zhongshan Hospital |  |  | Shidong Hospital | CodiEsp |
| --- | --- | --- | --- | --- | --- |
|  | Training | Development | Internal test | External test | Test |
| *Dataset and record characteristics* |  |  |  |  |  |
| Hospital stays or cases, n | 80,340 | 5,000 | 5,000 | 4,109 | 250 |
| Clinical setting | Inpatient | Inpatient | Inpatient | Inpatient | Curated clinical cases |
| Hospital level | Tertiary | Tertiary | Tertiary | Secondary | Not applicable |
| Language evaluated | Chinese | Chinese | Chinese | Chinese | Spanish |
| Diagnosis dictionary | Chinese ICD-10 based | Chinese ICD-10 based | Chinese ICD-10 based | Chinese ICD-10 based | CIE-10-ES |
| Notes per stay or case, median (IQR) | 7 (6–9) | 7 (6–9) | 7 (6–8) | 8 (6–9) | 1 (1–1) |
| Characters per stay or case, median (IQR) | 11,455<br>(9,371–14,163) | 11,520<br>(9,411.2–14,194.2) | 11,514.5<br>(9,371.8–14,291.8) | 5,426 (3,925–7,499) | 2,207<br>(1,544.75–2,960) |
| Diagnosis codes per stay or case, median (IQR) | 4 (3–6) | 4 (3–6) | 4 (2–6) | 5 (3–7) | 10 (6–15) |
| Unique diagnosis codes, n | 6,819 | 2,359 | 2,371 | 1,747 | 1,143 |
| *Age, sex, and length of stay* |  |  |  |  |  |
| Estimated age, years, median (IQR) | 65 (55–75) | 65 (55–75) | 65 (55–75) | 65 (65–75) | Not applicable |
| Age missing, n | 0 | 0 | 0 | 129 | Not applicable |
| Age < 20 years, n (%) | 962 (1.2%) | 55 (1.1%) | 55 (1.1%) | 63 (1.5%) | Not applicable |
| Age 20–39 years, n (%) | 8,930 (11.1%) | 565 (11.3%) | 583 (11.7%) | 190 (4.6%) | Not applicable |
| Age 40–59 years, n (%) | 26,734 (33.3%) | 1,688 (33.8%) | 1,658 (33.2%) | 627 (15.3%) | Not applicable |
| Age 60–79 years, n (%) | 40,482 (50.4%) | 2,482 (49.6%) | 2,488 (49.8%) | 2,496 (60.7%) | Not applicable |
| Age ≥ 80 years, n (%) | 3,232 (4.0%) | 210 (4.2%) | 216 (4.3%) | 604 (14.7%) | Not applicable |
| Age missing, n (%) | 0 | 0 | 0 | 129 (3.1%) | Not applicable |
| Male sex, n (%) | 46,602 (58.0%) | 2,941 (58.8%) | 2,911 (58.2%) | 1,885 (45.9%) | Not applicable |
| Female sex, n (%) | 33,738 (42.0%) | 2,059 (41.2%) | 2,089 (41.8%) | 2,224 (54.1%) | Not applicable |
| Sex missing, n | 0 | 0 | 0 | 0 | Not applicable |
| Length of stay, days, median (IQR) | 3 (2–5) | 3 (2–5) | 3 (2–5) | 6 (3–9) | Not applicable |
| Length of stay missing, n | 0 | 0 | 0 | 0 | Not applicable |
| *Detailed admission-department groups, n (%)* |  |  |  |  |  |
| Internal medicine | 28,736 (35.8%) | 1,769 (35.4%) | 1,795 (35.9%) | 1,795 (43.7%) | Not applicable |
| Surgery | 28,436 (35.4%) | 1,733 (34.7%) | 1,742 (34.8%) | 1,190 (29.0%) | Not applicable |
| Oncology or radiotherapy | 6,910 (8.6%) | 473 (9.5%) | 439 (8.8%) | 83 (2.0%) | Not applicable |
| Obstetrics and gynaecology | 2,545 (3.2%) | 149 (3.0%) | 160 (3.2%) | 282 (6.9%) | Not applicable |
| Paediatrics | 27 (<0.1%) | 1 (<0.1%) | 2 (<0.1%) | 173 (4.2%) | Not applicable |
| Ophthalmology, ENT, or stomatology | 3,024 (3.8%) | 198 (4.0%) | 206 (4.1%) | 147 (3.6%) | Not applicable |
| Critical care or emergency | 610 (0.8%) | 32 (0.6%) | 44 (0.9%) | 216 (5.3%) | Not applicable |
| Endoscopy or interventional services | 5,120 (6.4%) | 330 (6.6%) | 320 (6.4%) | 56 (1.4%) | Not applicable |
| Traditional Chinese medicine | 2,776 (3.5%) | 173 (3.5%) | 155 (3.1%) | 35 (0.9%) | Not applicable |
| Health management | 1,232 (1.5%) | 83 (1.7%) | 73 (1.5%) | 0 | Not applicable |
| Dermatology | 438 (0.5%) | 26 (0.5%) | 35 (0.7%) | 0 | Not applicable |
| Rehabilitation | 135 (0.2%) | 6 (0.1%) | 9 (0.2%) | 24 (0.6%) | Not applicable |
| Other | 351 (0.4%) | 27 (0.5%) | 20 (0.4%) | 0 | Not applicable |
| Missing | 0 | 0 | 0 | 108 (2.6%) | Not applicable |
| *Five most common original admission-department names, n (%)* |  |  |  |  |  |
| Rank 1 | Cardiology; 12,558 (15.6%) | Cardiology; 744 (14.9%) | Cardiology; 788 (15.8%) | Gastroenterology; 539 (13.1%) | Not applicable |
| Rank 2 | Endoscopy Centre; 4,707 (5.9%) | Endoscopy Centre; 297 (5.9%) | Endoscopy Centre; 292 (5.8%) | Cardiology; 357 (8.7%) | Not applicable |
| Rank 3 | Hepatobiliary Oncology and Liver Transplant Surgery; 4,386 (5.5%) | Hepatobiliary Oncology and Liver Transplant Surgery; 275 (5.5%) | Hepatobiliary Oncology and Liver Transplant Surgery; 264 (5.3%) | General Surgery Unit 1; 335 (8.2%) | Not applicable |
| Rank 4 | Thoracic Surgery; 4,071 (5.1%) | Thoracic Surgery; 263 (5.3%) | Thoracic Surgery; 257 (5.1%) | Neurology; 290 (7.1%) | Not applicable |
| Rank 5 | Haematology; 3,845 (4.8%) | Respiratory Medicine; 230 (4.6%) | Respiratory Medicine; 252 (5.0%) | Gynaecology; 282 (6.9%) | Not applicable |

Data are n, n (%), or median (IQR). Percentages use all records in each split. Masked 10-year age intervals were represented by their midpoints; fully suppressed values were missing, and age groups were aligned with the masking. Sex was recorded as male or female in the source data. Length of stay was discharge date minus admission date; same-day stays were assigned 0 days. Admission departments were harmonised by rule-based mapping, with original names translated into English. Note counts included all documents, and character counts excluded leading and trailing whitespace. Each CodiEsp case comprised one Spanish text; its word-count median (IQR) was 330.5 (235.5–442.75), with words defined by whitespace. Quartiles used linear interpolation. ENT=ear, nose, and throat; ICD=International Classification of Diseases.

### Exploratory computational throughput analysis

appendix table A2 reports the tested hardware, serving, batching, and measurement configuration and the observed results. The measurement was intended to characterise batch-processing throughput under one fixed local deployment, not theoretical hardware capacity, individual-stay latency, or coder labour time.

**Table A2:**
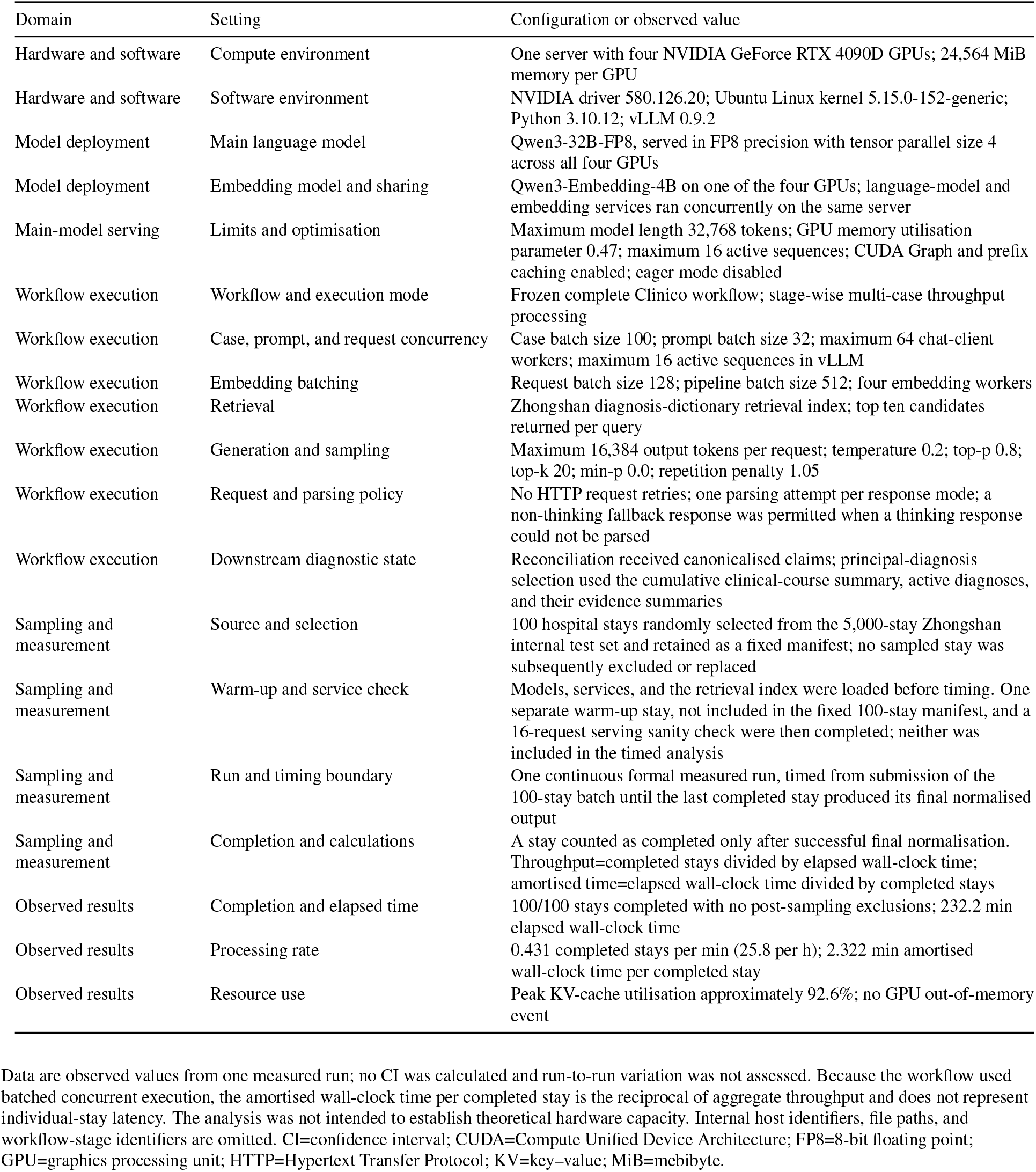
Configuration and results of the exploratory computational throughput analysis.

### Exploratory physician survey

The survey was administered separately from the retrospective performance evaluation, approximately 5 months after Clinico entered operational use. All items were required, the online platform limited each account to one submission, and all 64 submissions were complete. appendix table A3 presents unweighted item-level counts and percentages using all 64 voluntary respondents as the denominator, except for departmental coverage.

**Table A3:**
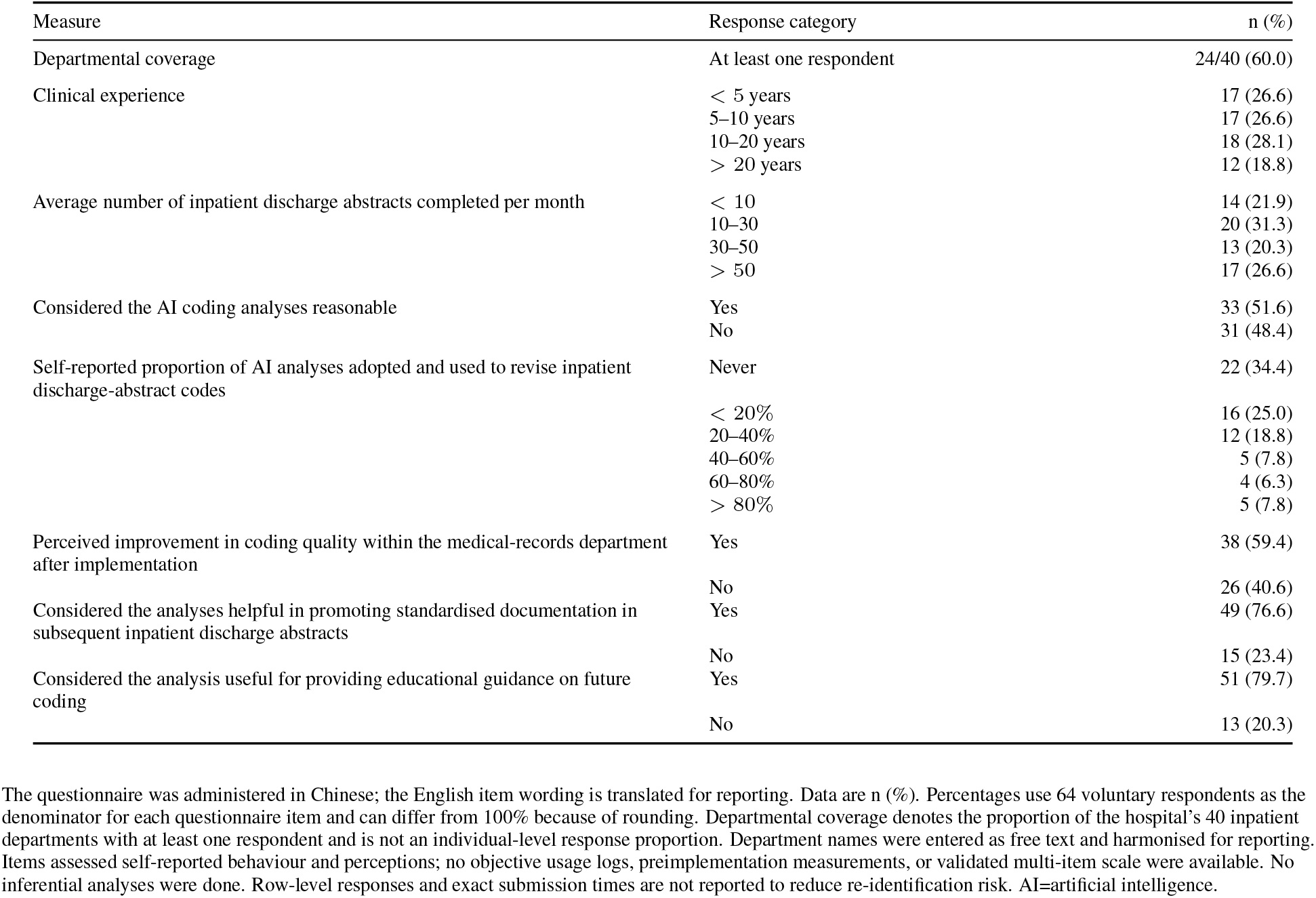
Respondent characteristics and responses in the exploratory physician survey.

### Post-hoc component ablation analysis

Complete results of the post-hoc component ablation analysis described in the main text are presented in appendix tables A4 and A5. Each ablation was initialised from the corresponding stored full-workflow state. Outputs upstream of the ablated component were retained; component-owned and dependent downstream outputs were removed, and the affected downstream stages were rerun once. Bootstrap resampling used the single stored output from each workflow configuration and therefore quantified hospital-stay sampling variation, not variability across repeated model generations.

**Table A4:**
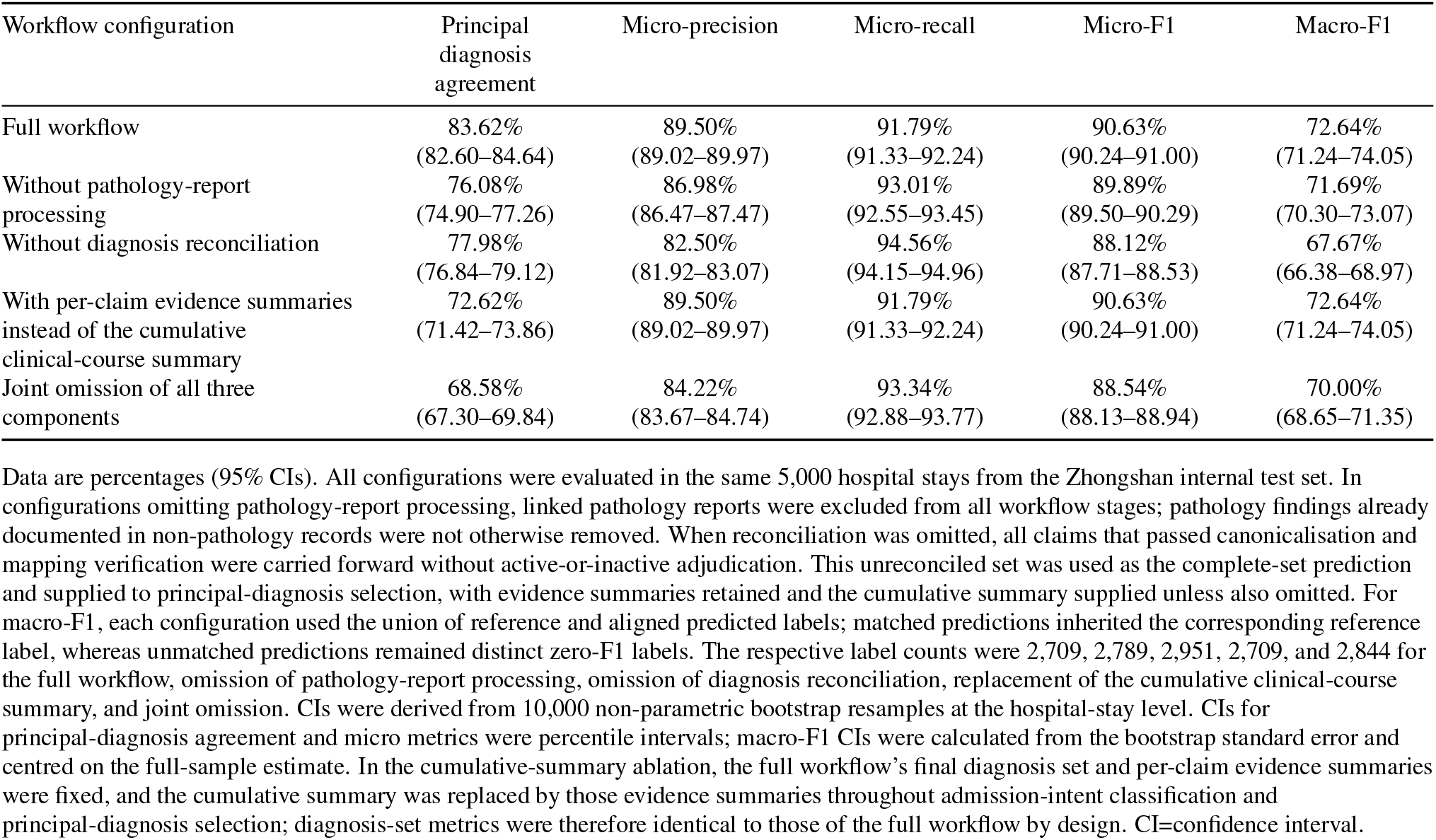
Performance of the full workflow and component-ablated configurations.

**Table A5:** Paired performance differences between the full workflow and component-ablated configurations.

| Comparison | $\Delta$ Principal diagnosis agreement | $\Delta$ Micro-precision | $\Delta$ Micro-recall | $\Delta$ Micro-F1 | $\Delta$ Macro-F1 |
| --- | --- | --- | --- | --- | --- |
| Full workflow vs without pathology-report processing | +7.54<br>(6.60–8.50) | +2.52<br>(2.20–2.83) | −1.22<br>(−1.50 to −0.94) | +0.73<br>(0.50–0.97) | +0.96<br>(0.51–1.41) |
| Full workflow vs without diagnosis reconciliation | +5.64<br>(4.82–6.48) | +6.99<br>(6.66–7.34) | −2.77<br>(−3.03 to −2.53) | +2.51<br>(2.27–2.74) | +4.97<br>(4.54–5.40) |
| Full workflow vs with per-claim evidence summaries instead of the cumulative clinical-course summary | +11.00<br>(9.92–12.04) | 0.00<br>(0.00–0.00) | 0.00<br>(0.00–0.00) | 0.00<br>(0.00–0.00) | 0.00<br>(0.00–0.00) |
| Full workflow vs joint omission | +15.04<br>(13.78–16.28) | +5.28<br>(4.93–5.63) | −1.55<br>(−1.83 to −1.27) | +2.09<br>(1.83–2.34) | +2.64<br>(2.19–3.10) |

Data are percentage-point differences (95% CIs), calculated as the full workflow minus the ablated workflow; positive values indicate higher performance with the full workflow. Differences were calculated from unrounded estimates. For paired macro-F1 differences, each configuration retained its own label union as defined in table A4. CIs were derived from 10,000 paired non-parametric bootstrap resamples at the hospital-stay level, with the same sampled hospital stays used for the full and corresponding ablated workflows in each resample. CIs for principal-diagnosis agreement and micro-metric differences were percentile intervals; macro-F1 difference CIs were calculated from the paired-bootstrap standard error and centred on the full-sample paired difference. In the cumulative-summary ablation, the final diagnosis set was identical to that of the full workflow by design. CI=confidence interval.

### Clinical examples of complex diagnostic evolution

These three cases were selected post hoc from de-identified study records. They are included to illustrate distinct diagnostic mechanisms, not to estimate their frequency or assess model performance. Exact dates and non-essential patient characteristics were omitted, and findings from the source clinical and pathology documents were paraphrased.

**Table A6:** Three de-identified hospital stays illustrating why chronology alone is insufficient.

| Diagnostic pattern | Evidence from the hospital record and diagnostic implication |
| --- | --- |
| Related thymic and metastatic diagnoses | An adult with a history of resected thymoma was admitted with suspected recurrence. Surgery identified a mediastinal mass and multiple pleural and left lower-lung nodules, but the discharge record retained the non-specific labels of mediastinal mass and postoperative thymic tumour. Separate specimens showed B3 thymoma invading extracapsular fat in the mediastinum, tumour metastasis in a pleural nodule, and multiple metastatic B3 thymomas in the lung. Chronology alone could not show that the specific thymic, pleural, and pulmonary diagnoses should coexist rather than replace one another. |
| Different specimens and lesions during liver transplantation | An adult admitted with cirrhosis and multiple indeterminate liver lesions underwent liver transplantation. Donor-liver biopsy showed mild steatosis and cholestasis, whereas examination of the explanted recipient liver confirmed three foci of moderately differentiated hepatocellular carcinoma. A thrombus removed from the recipient portal vein contained fibrous tissue and calcification, with no tumour reported, and later imaging showed an indeterminate nodule in the graft. Chronology alone was insufficient because these sequential findings concerned different specimens, organs, and lesions. |
| Different levels of certainty across tumour specimens | An adult inpatient underwent surgery for bilateral adnexal masses. Most same-day frozen sections showed at least borderline serous tumour without definite stromal invasion, whereas a right tubal specimen raised possible low-grade serous carcinoma; the discharge diagnosis therefore remained provisional. Permanent sections after discharge confirmed low-grade serous carcinoma in a fully sampled right ovarian specimen and cancer involvement in additional ovarian, tubal, omental, and peritoneal specimens. Because contemporaneous specimens differed in diagnostic certainty, the final diagnosis required integration of specimen-specific evidence rather than chronology alone. |

### Worked example of source-linked diagnosis adjudication

The first case in appendix table A6 was expanded to illustrate the workflow and is presented in abridged and de-identified form; it was not a separate validation analysis. Only diagnosis claims affecting the final diagnosis set are shown. Source-linked evidence and the cumulative hospital-course summary were shortened and paraphrased for presentation, without adding diagnostic information.

#### A. Initial diagnosis-claim discovery

| Initial diagnosis claim | Source and abridged evidence summary | Ledger action |
| --- | --- | --- |
| Postoperative thymic tumour | The record documented previous thymic-tumour resection. Admission, operative, and discharge documents continued to use a broad postoperative thymic-tumour diagnosis. | Added from admission documentation and subsequently confirmed. |
| Pleural metastatic tumour | Operative documentation described multiple pleural nodules and recorded pleural metastatic disease. | Added from operative documentation. |
| Mediastinal mass | Imaging and operative exploration identified a mediastinal lesion, while the discharge diagnosis remained non-specific. | Added from discharge documentation. |

No separate pulmonary secondary malignancy claim was identified from the non-pathology documents.

#### B. Pathology-derived diagnosis updates

| Specimen | Pathological evidence | Claim update |
| --- | --- | --- |
| Mediastinal mass | B3 thymoma with local invasion into extracapsular fat. | The broad thymic-tumour claim was refined to malignant neoplasm of the thymus. |
| Pleural nodule | Tumour metastasis was identified. | The pleural metastatic claim was confirmed and refined to secondary malignant neoplasm of the pleura. |
| Left lower-lung wedge | Multiple metastatic B3 thymomas were identified. | A new claim of secondary malignant neoplasm of the lung was added. |

Pathology review therefore had three different effects in this case: it refined a broad diagnosis, confirmed an existing metastatic claim, and added a diagnosis that had not been established from the non-pathology documents.

#### C. Diagnostic chains

After pathology review, Clinico organised the claims into three lesion-specific diagnostic chains (appendix figure A1).

**Figure A1:**
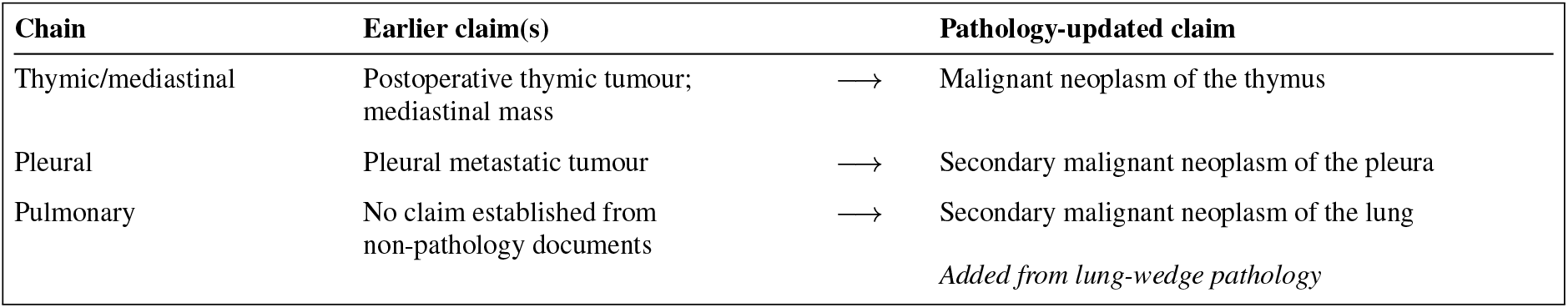
Diagnostic chains in the worked example. Pathology refined the thymic diagnosis, confirmed the pleural metastatic diagnosis, and added a separate pulmonary metastatic diagnosis.

#### D. Diagnosis reconciliation

##### Active diagnoses

Malignant neoplasm of the thymus; secondary malignant neoplasm of the pleura; secondary malignant neoplasm of the lung.

##### Inactive diagnoses

Postoperative thymic tumour, superseded by the more specific pathology-supported thymic diagnosis; mediastinal mass, a non-specific anatomical description covered by the pathology-supported thymic diagnosis.

The thymic, pleural, and pulmonary diagnoses were related but not redundant. Because they represented different anatomical sites, all three specific diagnoses remained active.

#### E. Cumulative hospital-course summary and final output

##### Cumulative hospital-course summary (abridged)

The record documented previous resection of a thymic tumour. The patient was admitted with suspected recurrent thoracic disease after follow-up imaging showed mediastinal, pleural, and pulmonary abnormalities. Operative exploration identified a mediastinal mass and multiple pleural and left lower-lung nodules, from which specimens were obtained. Admission and discharge documentation continued to use the broad labels of postoperative thymic tumour and mediastinal mass. Linked pathology showed B3 thymoma with extracapsular fat invasion in the mediastinal specimen, tumour metastasis in a pleural nodule, and multiple metastatic B3 thymomas in the lung wedge. Taken together, the record supported an invasive thymic neoplasm with separate pleural and pulmonary metastases.

##### Final diagnosis-code set

Malignant neoplasm of the thymus (C37.×00); secondary malignant neoplasm of the pleura (C78.200); secondary malignant neoplasm of the lung (C78.000×011).

##### Principal diagnosis

Secondary malignant neoplasm of the pleura (C78.200).

